# Evaluating the Effectiveness and Safety of Large Language Model in Generating Type 2 Diabetes Mellitus Management Plans: A Comparative Study with Medical Experts Based on Real Patient Records

**DOI:** 10.1101/2024.05.21.24307700

**Authors:** Agnibho Mondal, Arindam Naskar, Bhaskar Roy Choudhury, Sambudhya Chakraborty, Tanmay Biswas, Sumanta Sinha

## Abstract

**Background:** The integration of large language models (LLMs) such as GPT-4 into healthcare presents potential benefits and challenges. While LLMs have shown promise in applications ranging from scientific writing to personalized medicine, their practical utility and safety in clinical settings remain under scrutiny. Concerns about accuracy, ethical considerations and bias necessitate rigorous evaluation of these technologies against established medical standards.

**Objective:** To compare the completeness, necessity, dosage accuracy and overall safety of type 2 diabetes management plans created by GPT-4 with those devised by medical experts.

**Methods:** This study involved a comparative analysis using anonymized patient records from a healthcare setting in West Bengal, India. Management plans for 50 Type 2 diabetes patients were generated by GPT-4 and three blinded medical experts. These plans were evaluated against a reference management plan based on American Diabetes Society guidelines. Completeness, necessity and dosage accuracy were quantified and an error score was devised to assess the quality of the generated management plans. The safety of the management plans generated by GPT-4 was also assessed.

**Results:** Results indicated that medical experts’ management plans had fewer missing medications compared to those generated by GPT-4 (p=0.008). However, GPT-4 generated management plans included fewer unnecessary medications (p=0.003). No significant difference was observed in the accuracy of drug dosages (p=0.975). The overall error scores were comparable between human experts and GPT-4 (p=0.301). Safety issues were noted in 16% of the plans generated by GPT-4, highlighting potential risks associated with AI-generated management plans.

**Conclusion:** The study demonstrates that while GPT-4 can effectively reduce unnecessary drug prescriptions, it does not yet match the performance of medical experts in terms of plan completeness and safety. The findings support the use of LLMs as supplementary tools in healthcare, underscoring the need for enhanced algorithms and continuous human oversight to ensure the efficacy and safety of AI applications in clinical settings. Further research is necessary to improve the integration of LLMs into complex healthcare environments.

## Introduction

As the frontier of artificial intelligence (AI) continues to advance, the integration of large language models (LLMs) such as GPT-4 into healthcare settings presents both promising opportunities and significant challenges. The potential of LLMs to enhance healthcare education, research and practice is noteworthy, with applications ranging from improving scientific writing to assisting in complex data analysis and personalized medicine. However, the deployment of these technologies in clinical environments must be approached with caution due to concerns about accuracy, ethical considerations and the potential for bias.[1]

LLMs like GPT-4[2] have shown significant potential in a range of healthcare applications, from generating patient management plans to assisting with medical documentation. Despite their capabilities, the accuracy and utility of these models in practical, clinical settings require thorough evaluation and benchmarking against established medical standards. Studies have noted that while LLMs perform well in tasks like answering medical exam questions, their application in direct patient care and other complex medical scenarios remains underexplored and often lacks integration with real patient data.[3]

Moreover, the evaluation of these models in healthcare has often focused narrowly on specific tasks, such as NLP tasks related to summarization and conversation, without a broad application across various medical specialties. This has limited the understanding of their broader potential and areas where they may not perform as expected. To truly harness the capabilities of LLMs like GPT-4, comprehensive assessments using real-world data and across diverse healthcare tasks are essential.[1]

The advanced capabilities of LLMs may extend beyond routine natural language processing to include complex clinical interactions and decision-making processes. It is necessary to carry out more accurate assessment of LLMs’ functionality in real-world clinical scenarios.[4]

Recent studies highlight the escalating global diabetes burden, advocating for innovative management strategies like the application of large language models (LLMs), including GPT-4. These models offer promising advances in diabetes care by potentially enhancing guideline adherence and providing personalized, evidence-based treatment recommendations. Research involving a comparative analysis of hypothetical diabetes cases assessed by GPT-4 against expert evaluations indicates a high concordance, suggesting LLMs’ capability to support healthcare professionals in diabetes management. However, discrepancies in complex clinical judgments call for further refinement of AI technologies.[5]

This study aims to fill these gaps by directly comparing the management plans created by GPT-4 with those devised by medical experts, focusing on Type 2 diabetes—a prevalent and complex medical condition. The comparison will consider several dimensions of evaluation, including the accuracy, comprehensiveness and practical usability of the management plans, providing a clearer picture of where LLMs stand in terms of replacing or augmenting traditional healthcare processes.

By addressing these critical points, the article will contribute valuable insights into the current capabilities and future potential of LLMs in healthcare, informing both technological developers and healthcare professionals about the strengths and limitations of these advanced AI tools in managing chronic diseases like Type 2 diabetes.

## Methodology

This comparative study evaluated the performance of GPT-4, against management plans created by medical experts for Type 2 diabetes. The study was designed to compare key metrics in three domains including completeness, necessity and accuracy.

The study involved anonymized patient records of type 2 diabetes mellitus patients from a healthcare database from the private practice of the second author in West Bengal, India. These records include comprehensive patient data including case summaries, laboratory reports and medication history.

Management plans for each of the selected patient records were generated through both GPT-4 and the medical experts. Three medical experts created the management plans independently who were blinded to the responses generated by each other. The application programming interface (API) of GPT-4 was used for generating the management plans.

A reference management plan was carefully developed in collaboration by the first two authors using the American Diabetes Society (ADA) guidelines.[6] The management plans generated by the medical experts and GPT-4 were evaluated against this reference.

As there was no preexisting metric suitable for such a comparative study between large language model and medical experts, we devised an error scoring for this purpose as follows:

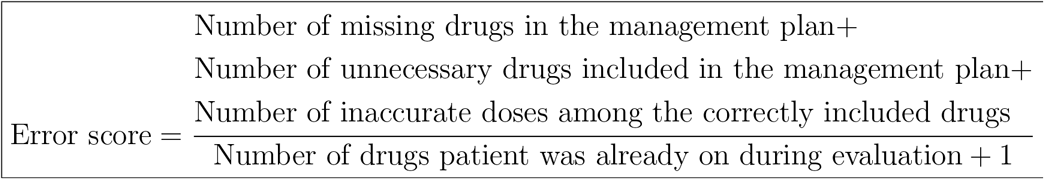

The number of missing drugs, number of unnecessary drugs and the number of inaccurate doses was determined based on the comparison with the reference management plan.

Management plans generated by each medical expert as well as the one generated by GPT-4 was evaluated separately against the reference by the third author who did not participate in the preparation of the reference management plans. Each of the 50 management plans were scored separately for each expert and GPT-4. The error scores of the three medical experts were then averaged for each management plan and the average error scores were used for comparison with the error scores obtained by GPT-4.

The methodology is shown as a flowchart in figure 1.

**Figure 1:**
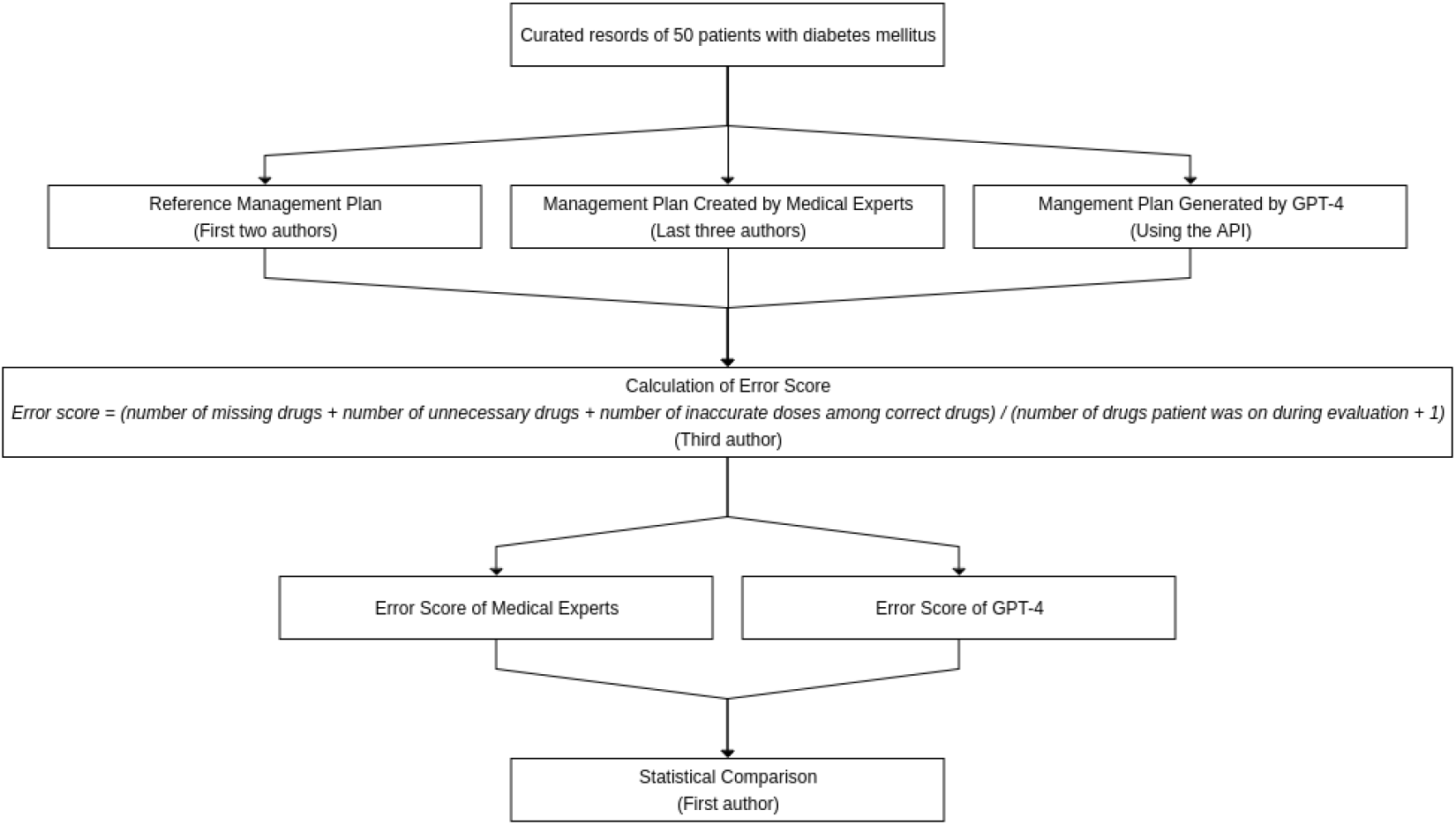
Flowchart showing the methodology

In addition, the management plans generated by GPT-4 were separately evaluated for potential safety issues. However, it was not included in the comparison.

Statistical analysis conducted using R Software by R Foundation for Statistical Analysis. The primary analysis involved comparison of average error scores obtained by the medical experts and those obtained by GPT-4. The error scores were described with mean and standard deviation (SD). Wilcoxon rank sum test was used for the comparison. The effect size was shown with a 95% confidence interval (95% CI). A p value of less than 0.05 was considered significant.

The study protocol was reviewed and approved by Clinimed Independent Ethics Committee, Kolkata with the reference number CLPL/CIEC/001/2024. All case records were deanonymized prior to inclusion in the study and data handling procedures strictly followed ethical guidelines.

## Results

This study compared the management plans for 50 type 2 diabetes mellitus patient records generated by medical experts and GPT-4 against a reference plan in accordance with the ADA guidelines.

### Completeness

the mean number of missing drugs in the management plan generated by the medical experts was 1.15 (SD 0.8). In comparison, the mean number of missing drugs in GPT-4 generated management plan was 1.76 (SD 1.2). The difference between these two was significant (p=0.008).

### Necessity

the mean number of unnecessary drugs in the management plan created by the medical experts was 0.68 (SD 0.64) while it was 0.4 (SD 0.61) in the management plan generated by GPT-4. The difference was significant (p=0.003).

### Dose accuracy

The mean number of inaccurate dosages of correctly included drugs was 0.74 (SD 0.58) in management plans generated by the medical experts and 0.86 (SD 0.9). The difference was not significant (p=0.975).

### Error score

The mean error score of the management plans formulated by the medical experts was 0.41 (SD 0.23) while it was 0.46 (SD 0.25) for GPT-4. A boxplot showing the comparison is shown in figure 2. There was no significant difference between the medical experts and GPT-4 in this regard (p=0.301). The difference in location was -0.05 (95% CI -0.14 to 0.05).

**Figure 2:**
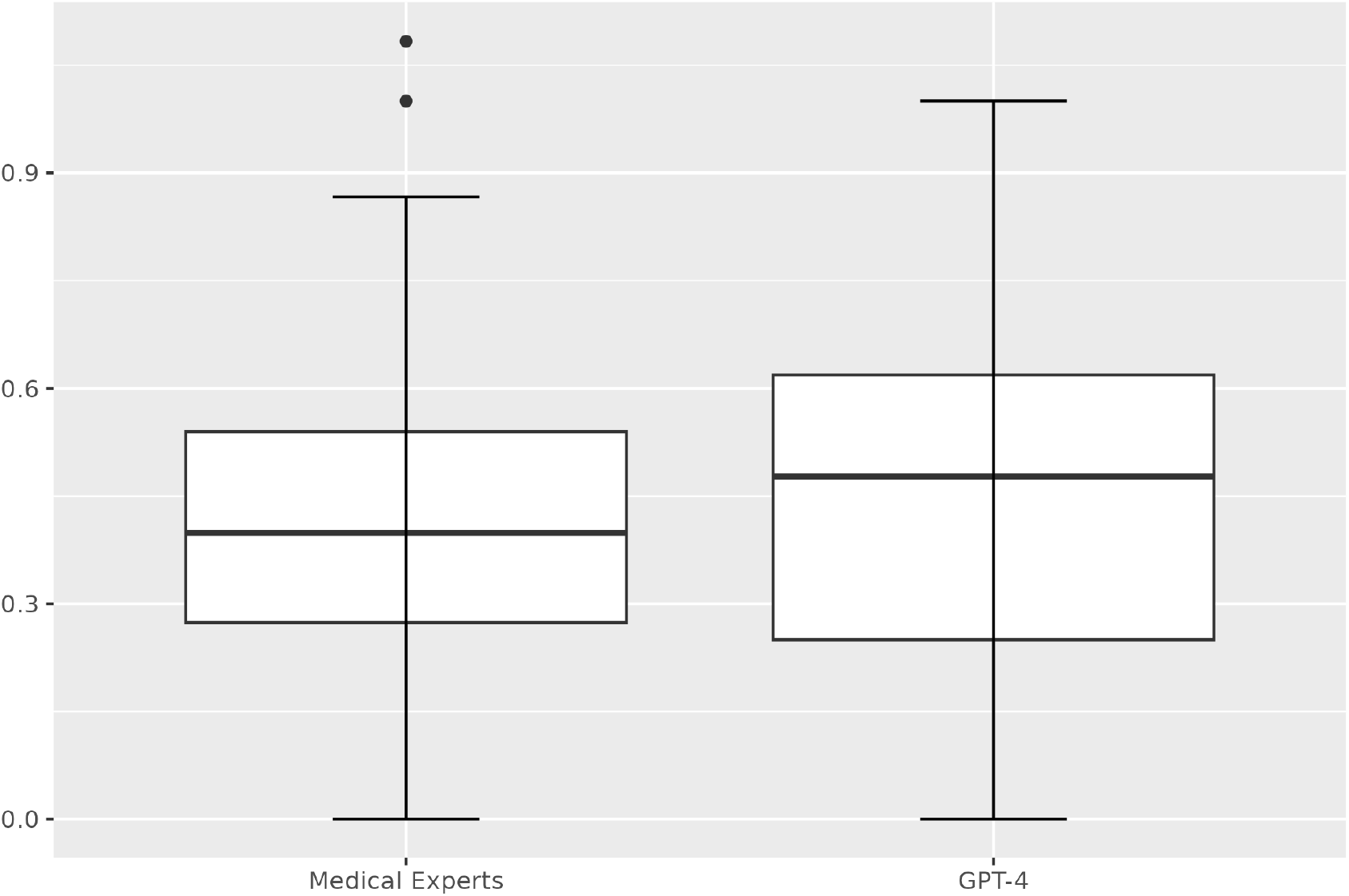
Boxplot comparing error scores of medical experts and GPT-4

The findings of the study are shown in table 1.

**Table 1:** Comparison of Medical Experts and GPT-4.

|  | Medical Experts<sup>*†</sup> | GPT-4<sup>†</sup> | p-value | Difference in location | 95% CI |
| --- | --- | --- | --- | --- | --- |
| Number of missing drugs | $1.15 \pm 0.8$ | $1.76 \pm 1.2$ | 0.008 | -0.67 | -1.0 to -0.33 |
| Number of unnecessary drugs included | $0.68 \pm 0.64$ | $0.4 \pm 0.61$ | 0.003 | 0.33 | 0 to 0.67 |
| Number of inaccurate doses among the correctly included drugs | $0.74 \pm 0.58$ | $0.86 \pm 0.9$ | 0.975 | 0 | -0.33 to 0.33 |
| Error score | $0.41 \pm 0.23$ | $0.46 \pm 0.25$ | 0.301 | -0.05 | -0.14 to 0.05 |

\* Averaged score of three medical experts
† Mean ± standard deviation

### Safety

there were safety issues in eight (16%) management plans generated by GPT-4. Five of them were due to an elevated risk of hypoglycemia and three were due to safety concerns with the drugs used. In one case GPT-4 did not withhold SGLT2 inhibitor despite recurrent urinary tract infection. In another case GPT-4 continued saxagliptin despite congestive heart failure. In another case GPT-4 increased dose of atorvastatin despite raised liver enzymes.

### Referral

In the management plans generated by GPT-4, the referrals were generally appropriate and comprehensive including referrals to ophthalmologists, nephrologists and other specialties. In only one case (2%) GPT-4 missed to refer the case to nephrologist despite deteriorating renal function.

## Discussion

The findings from this study reveal significant insights into the application of LLMs, specifically GPT-4, in generating management plans for type 2 diabetes mellitus compared to traditional plans formulated by medical experts. Our analysis focused on four key aspects: completeness, necessity, dose accuracy and safety of the management plans.

Our results indicate that management plans generated by medical experts were more complete than those produced by GPT-4, with fewer missing drugs on average (1.15 vs. 1.76, p=0.008). This suggests that while GPT-4 can generate management plans, it may occasionally omit necessary medications, potentially affecting the overall effectiveness of the treatment. The higher standard deviation in the GPT-4 group indicates a broader variability in the completeness of the AI-generated plans. Enhancements in the training algorithms of AI systems could potentially address these shortcomings, ensuring that all necessary medications are consistently included in the plans.

Conversely, the necessity of the drugs included in the management plans shows an interesting trend. GPT-4 tended to include fewer unnecessary medications compared to medical experts (0.4 vs. 0.68, p=0.003). This indicates a potential strength of AI in identifying and adhering more strictly to the most relevant treatment protocols, possibly by leveraging large datasets to determine the most commonly effective medications without as much bias or variability as human prescribers.

The accuracy of dosages for correctly included drugs did not differ significantly between the groups (p=0.975). Both medical experts and GPT-4 showed similar capabilities in dosing accuracy, suggesting that once a drug is identified as necessary, AI systems are reasonably effective in prescribing appropriate dosages. This finding supports the use of AI as a supportive tool in managing complex conditions like diabetes, where dosage precision is critical.

The overall error scores between the two groups were also statistically insignificant (p=0.301), further demonstrating the potential of AI in managing type 2 diabetes. The similarity in error scores suggests that GPT-4 can perform at a level comparable to medical experts in terms of overall plan quality.

However, a significant concern highlighted by our study is the safety of AI-generated plants. Safety issues were identified in 16% of the plans generated by GPT-4. These included risks of hypoglycemia and inappropriate continuation of medications in the presence of contraindications like recurrent urinary tract infection, congestive heart failure and raised liver enzymes. Such findings underline the critical need for improved algorithms that can recognize and integrate complex clinical scenarios and patient histories into their decision-making processes. It also demonstrates the importance of human oversight in preventing safety concerns associated with the use of AI in healthcare.

We observed high accuracy of GPT-4 in generating appropriate and comprehensive referrals which demonstrates its potential utility in supporting healthcare professionals by streamlining patient management and facilitating timely specialist consultations. However, the occasional oversight, such as the missed referral to a nephrologist highlights the need for continued oversight and regular evaluation of AI-generated recommendations.

Regarding implementation of technological innovations in healthcare, it has been observed that technological interventions often perform comparably to traditional methods in controlled settings but may vary in real-world applications.[7] To evaluate how well LLM would perform in real-world scenarios, we compared its performance against that of medical experts using actual patient records.

Recent advancements in diabetes management have emphasized a more nuanced approach, focusing not just on controlling blood sugar but also on preventing complications and managing the disease’s broad systemic impacts.[8] To successfully integrate LLMs into the complex field of therapy, it’s essential that these models do more than just replicate the guideline-based recommendations of healthcare professionals. They should also be capable of supporting complex decision-making that is tailored to the specific contexts of individual patients.

Furthermore, despite extensive research and the implementation of various new treatment protocols, including the increasing use of GLP-1 receptor agonists and SGLT2 inhibitors due to their benefits beyond glycemic control, management of Type 2 diabetes remains a complex challenge that requires a multifaceted approach.[8] This complexity might explain why automated systems like GPT-4, though proficient within the scope of their training, do not yet surpass human practitioners in terms of clinical decision-making and personalized patient care.

Moreover, the chronic care model and its emphasis on systematic, patient-centered care highlight the importance of integrating community resources and sustained lifestyle interventions, areas where human caregivers currently have a more significant impact compared to AI.[7] As such, while LLMs can support healthcare providers by offering data-driven insights and freeing up time from administrative tasks, the holistic management of chronic diseases like Type 2 diabetes still benefits significantly from the personal touch and clinical acumen of human practitioners.[9]

Overall, the study’s outcome emphasizes the potential of LLMs as supportive tools rather than replacements in healthcare, suggesting that further research and development are needed to enhance their practical application in clinical settings. Future studies should continue to explore how these technologies can be best utilized to complement the evolving paradigms in diabetes care, particularly in managing complex cases where lifestyle, medication and patient education are interlinked.

This study, while insightful, carries several limitations that should be considered when interpreting the results and planning future research. The study was based on a relatively small sample of 50 patient records. This limited sample size might not capture the full spectrum of variability in type 2 diabetes management across different populations or healthcare settings. The evaluation of management plans against ADA guidelines was performed by human experts, which introduces potential for subjective bias in interpreting the guidelines or the completeness and necessity of medications. We tried to limit this bias by preparing a reference management plan beforehand and comparing the generated management plans against it. Future studies might benefit from employing a standardized, automated scoring system to reduce this potential bias. AI technologies, including LLMs, are rapidly evolving. The findings from this study may not necessarily hold in the future as newer versions or different AI systems could exhibit different levels of effectiveness or safety in clinical practice.

Building on the findings and limitations of the current study, future research should be directed towards several key areas to enhance the understanding and application of AI in managing type 2 diabetes and other complex chronic conditions. Future studies should aim to include a larger and more diverse cohort of patient records from multiple healthcare settings worldwide. This would allow for a broader evaluation of AI capabilities across varied demographic and clinical contexts, providing a more robust assessment of its generalizability and effectiveness.

Investigating the impact of different training data sets on AI performance is essential as well. Studies should focus on training AI systems with data that include a wide range of patient scenarios, including those with multiple comorbidities, to understand how AI handles complex clinical situations. Moreover, the impact of real-time learning and adaptation in AI algorithms could be explored to see how these systems evolve with ongoing use in clinical environments.

Implementing automated systems for evaluating AI-generated management plans against clinical guidelines could reduce human bias and increase the reproducibility of results. Research should focus on developing and validating such automated systems, which could also be used in real-time clinical decision support.

Longitudinal studies that track the outcomes of patients managed with the assistance of AI over time would provide invaluable insights into the long-term effectiveness and safety of AI-supported treatment plans. Additionally, pilot studies that implement these AI systems in real-world clinical settings could evaluate practical challenges and patient outcomes in a routine clinical context.

As AI becomes more integrated into patient care, it is imperative to conduct research on the ethical implications and develop robust regulatory frameworks to ensure patient safety and data privacy. Studies should also examine patients’ and healthcare providers’ perceptions of and trust in AI technologies to identify barriers to adoption.

Comparing the performance of different AI models, including newer versions of GPT and other AI platforms, in the same tasks can help determine which models are most effective for specific aspects of diabetes management. This could guide healthcare providers in selecting the most appropriate AI tools for their needs.

These future studies would not only address the gaps identified in this research but also advance the field towards more effective, personalized and AI-integrated healthcare solutions.

## Conclusion

This study provides valuable insights into the utility and limitations of using LLMs, in the management of type 2 diabetes mellitus. The findings indicate that while AI can effectively minimize unnecessary drug prescriptions, it still lags behind medical experts in terms of the completeness and safety of management plans. Although AI demonstrates potential in matching human performance in some respects like dosage accuracy and overall error scores, significant concerns remain regarding safety, particularly in managing complex patient conditions. These results underscore the importance of human oversight in reviewing AI-generated treatment plans, especially in nuanced clinical scenarios.

## Data Availability

All data produced in the present study are available upon reasonable request to the authors

## Disclosure on writing assistance

Writing assistance was obtained from GPT-4. However, the authors are solely responsible for the content of the article.

